# Study Protocol Efficacy, safety, and tolerability of ultra-high-caloric, fatty diet (UFD) in amyotrophic lateral sclerosis

**DOI:** 10.1101/2024.11.21.24311161

**Authors:** Johannes Dorst, Christian Ruckes

**Affiliations:** University Hospital Ulm, Department of Neurology, University of Ulm, Oberer Eselsberg 45, D-89081 Ulm / Germany; Interdisciplinary Center for Clinical Trials (IZKS), University Medical Center of the Johannes-Gutenberg, University Mainz, Langenbeckstraße 1, 55131 Mainz

## Abstract

The LIPCAL-ALS II study investigates the efficacy, safety, and tolerability of an ultra-high-caloric, fatty diet (UFD) in amyotrophic lateral sclerosis (ALS). It is a multicenter, 1:1-randomized, prospective, stratified, parallel-group, double-blind, placebo-controlled, confirmatory study with 392 patients conducted in 25 centers of the German network for motor neuron diseases (MND Net). Patients will be randomized to receive either the experimental intervention, consisting of an ultra-high-caloric fatty dietary supplement for drinking (100% lipid, 4.5 kcal/ml) 140 ml/day (630 kcal) or matching placebo (140 ml/day, <5% fat, <50 kcal) in addition to normal food intake. The primary endpoint is survival after 18 months of intervention. Patients with possible, probable (clinically or laboratory supported) or definite ALS according to revised El Escorial criteria with a disease duration of <36 months and a loss of ALS functional rating scale revised (ALSFRS-R) ≥0.33 points/months will be included.

**Synopsis:** 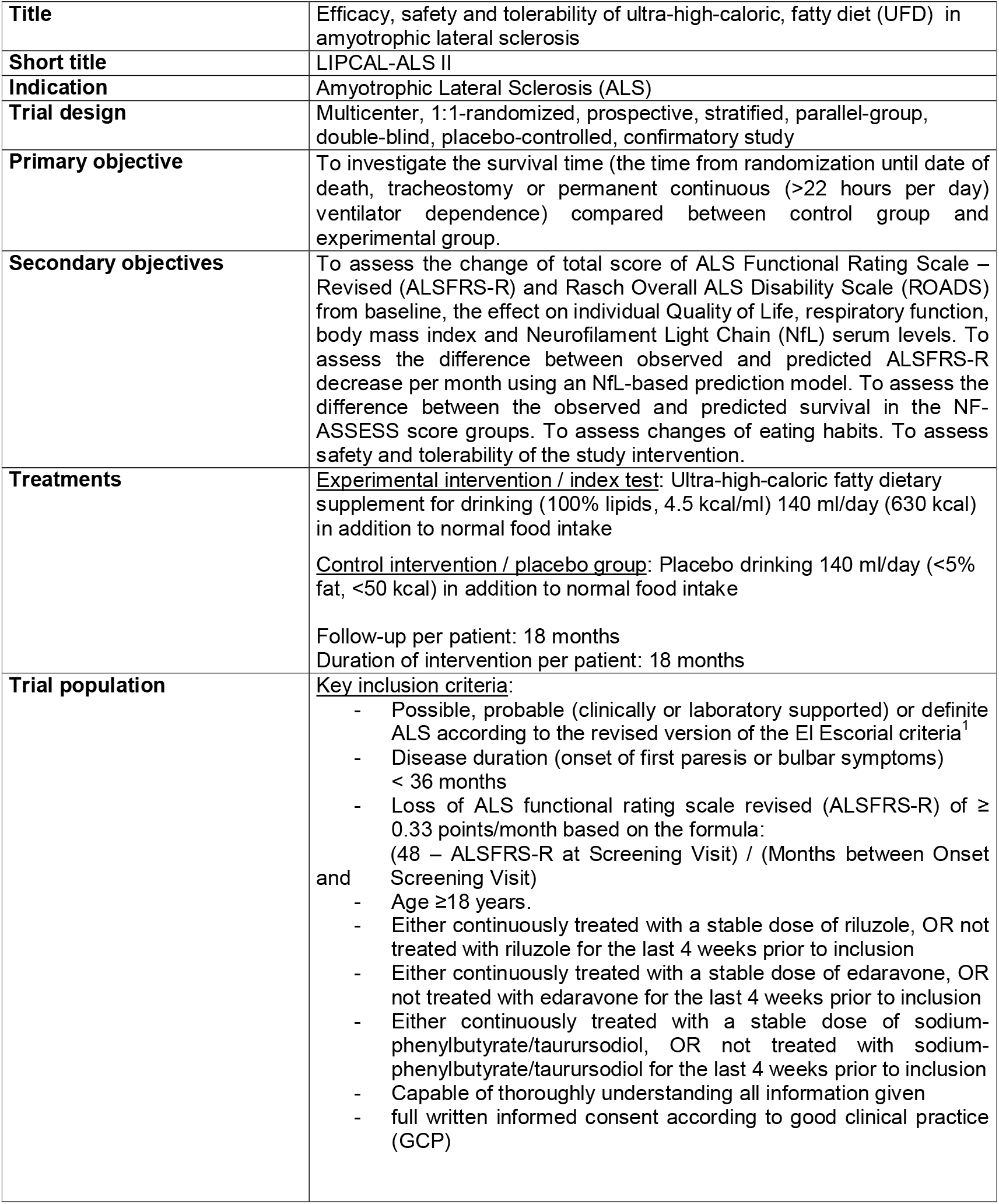

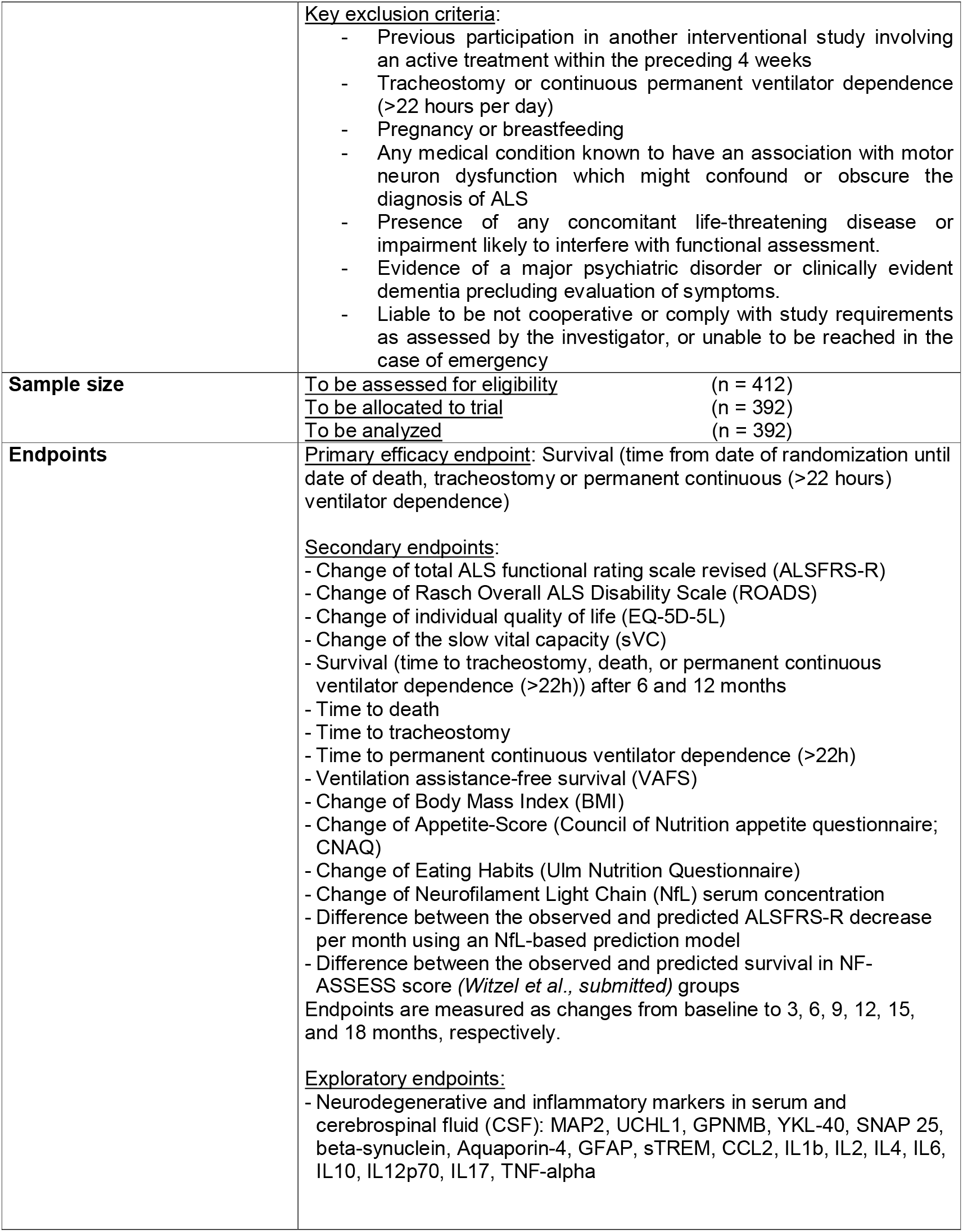

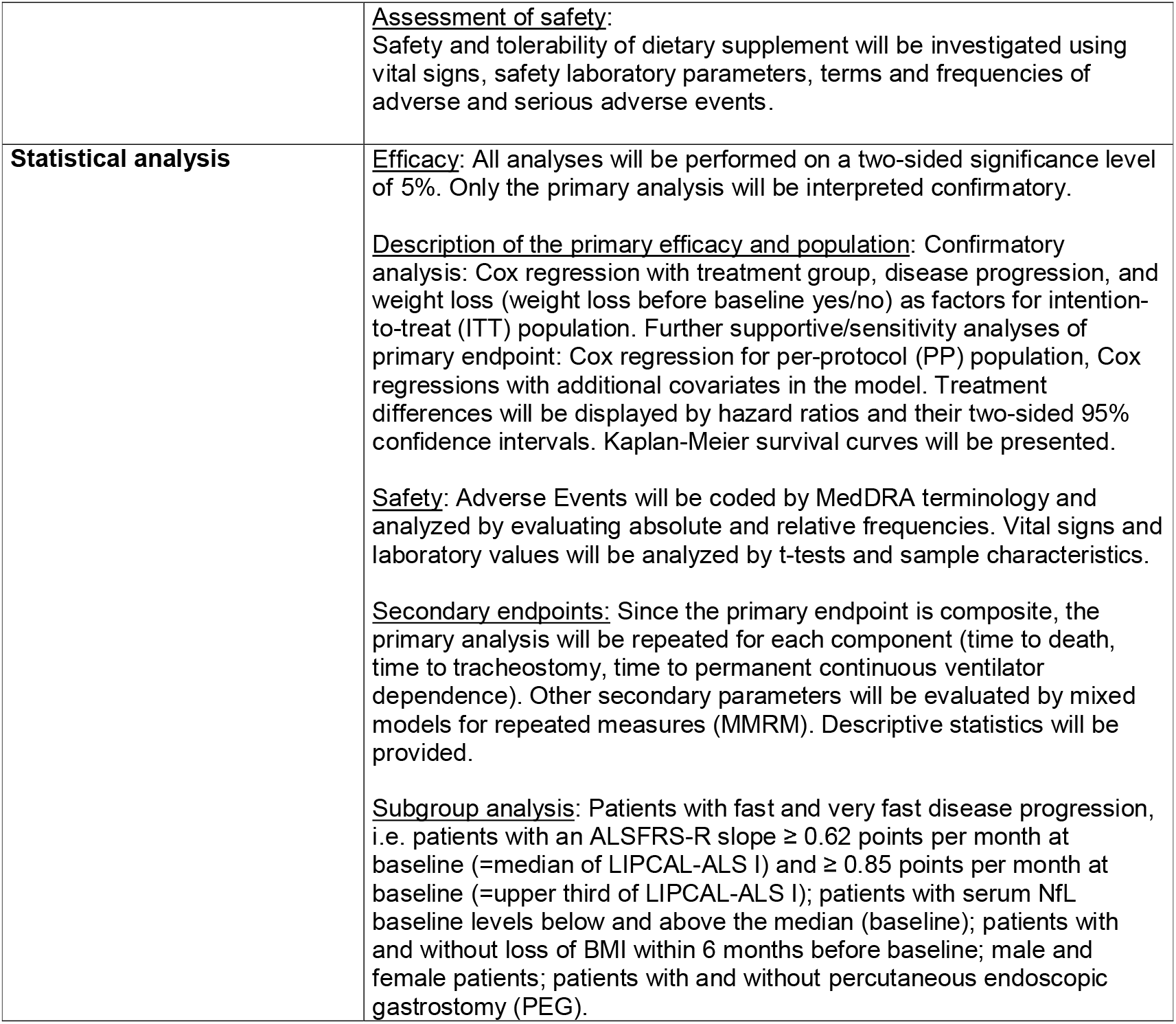

## Trial schedule

**Table 1:** Schedule of study procedures:

|  | V1<br>On-Site<br>(SCR/BL) | T1<br>Phone | T2<br>Phone | T3<br>Phone | V2<br>On-Site | V3<br>On-Site | T4<br>Phone | V4<br>On-Site | T5<br>Phone | V5<br>On-Site |
| --- | --- | --- | --- | --- | --- | --- | --- | --- | --- | --- |
|  | 0 | 5d | 2w | 1m | 3m | 6m | 9m | 12m | 15m | 18m |
| Informed consent | X |  |  |  |  |  |  |  |  |  |
| Inclusion/Exclusion Criteria | X |  |  |  |  |  |  |  |  |  |
| Medical History | X |  |  |  |  |  |  |  |  |  |
| Concomitant Treatment | X |  | X | X | X | X | X | X | X | X |
| Diagnostic Criteria<sup>a</sup> | X |  |  |  |  |  |  |  |  |  |
| Physical Examination<sup>b</sup> | X |  |  |  | X | X |  | X |  | X |
| Laboratory Blood Tests<sup>c</sup> | X |  |  |  | X | X |  | X |  | X |
| Pregnancy Test | X |  |  |  |  |  |  |  |  |  |
| Lumbar puncture (voluntary) | (X) |  |  |  |  | (X) |  | (X) |  | (X) |
| ALSFRS-R<sup>d</sup> | X |  |  |  | X | X | X | X | X | X |
| ROADS<sup>e</sup> | X |  |  |  | X | X | X | X | X | X |
| SVC<sup>f</sup> | X |  |  |  | X | X |  | X |  | X |
| EQ-5D-5L<sup>g</sup> | X |  |  |  | X | X |  | X |  | X |
| CNAQ<sup>h</sup> | X |  | X | X | X | X | X | X | X | X |
| Ulm Nutrition Questionnaire<sup>i</sup> | X |  | X | X | X | X | X | X | X | X |
| Ulmer Fragebogen zur Studienzufriedenheit<sup>j</sup> | X |  |  |  | X | X |  | X |  | X |
| Adverse events<sup>k</sup> |  | X | X | X | X | X | X | X | X | X |
| Tolerability and Compliance |  | X | X | X | X | X | X | X | X | X |

\*) combined screening and baseline visit
a) Acquisition of ALS diagnostic criteria (revised El Escorial criteria<sup>1</sup>), familial history, site of onset (spinal or bulbar), development of body weight during the last 6 months preceding baseline
b) vital signs, height, weight, blood pressure, heart rate, internal and neurological examination
c) blood count, electrolytes, liver and kidney values, CRP, glucose, Hba1c, cholesterol, HDL, LDL, triglycerides, CK, NFL, exploratory/scientific parameters
d) ALS Functional Rating Scale Revised (ALSFRS-R<sup>2</sup>) and progression rate
e) Rasch-Built Overall Amyotrophic Lateral Sclerosis Disability Score (ROADS<sup>3</sup>)
f) Slow vital capacity (SVC; best of 3 measurements)
g) Evaluation of quality of life (Euro Quality of Life-5D-5L, EQ-5D-5L)
h) Evaluation of appetite (Council of Nutrition Appetite Questionnaire; CNAQ)
i) Evaluation of nutrition habits (Ulm Nutrition Questionnaire) and one question whether patients believed that they were receiving UHCFD or placebo
j) Evaluation of study contentment, study-related quality of life, and personal motivation for study participation (new score, which shall be validated during this study)
k) Evaluation of Adverse Events (AEs) and Serious Adverse Events (SAEs) by anamnesis, physical examination, and safety laboratory parameters

## 1 INTRODUCTION

### 1.1 Scientific background / Trial Rationale

ALS is a fatal neurodegenerative disease, leading to progressive paralysis of voluntarily innervated muscles and to death caused by respiratory failure after a mean disease duration of 2–4 years^4^. The incidence of ALS in Europe is 2.1–3.8/100.000 person-years^4^, close to the incidence of Multiple Sclerosis^5^. At the same time, caused by the extremely low life expectancy, prevalence is only 4.1– 8.4/100.000 persons^4^. The age-adjusted mortality rate was 1.7 between 2011–2014 in the US^6^. Considering various metrics of disease burden (years of life lost (YLLs), years of life lived with disability (YLDs), disability-adjusted life-years (DALYs)) the global burden of the disease is very high^7^. Furthermore, the burden of ALS has increased substantially over the last 20 years^7^.

Despite decades of intensive research, the pathogenesis of ALS remains largely unknown. It has been shown that p-TDP-43, a pathologically misfolded protein, continuously spreads through the brain and spinal cord^8,9^. However, so far, this finding has not been translated to an effective therapy. To date, riluzole, a glutamate antagonist, is the only approved drug for ALS in Europe, marginally extending survival by about 3 months based on a trial from 1996^10^. Since then, despite over 25 years of intensive research, no further effective treatments have been identified, and promising therapeutic approaches remain scarce. Edaravone, a free radical scavenger, and sodium phenylbutyrate-taurursodiol, a combination of two neuroprotective substances, were approved in the US after each showing a rather small effect on disease progression^11^. In Europe, edaravone is still in clinical development as has not been approved so far. Regarding sodium phenylbutyrate-taurursodiol, EMA awaits the results of a second, larger pivotal study, which are expected in the second half of 2024. Ongoing studies with antisense oligonucleotides (ASOs) are very promising^12,13^, but are only applicable for genetically determined ALS forms (∼10% of total cases). Thus, we can conclude that riluzole still constitutes the only approved drug and the standard therapy in Europe, and that more effective treatments for ALS are desperately needed.

The proposed study aims at improving survival of ALS patients by targeting metabolic parameters. ALS patients feature an intrinsic hypermetabolism as signified by an increased resting energy expenditure^14^ which (among other factors) significantly contributes to progressive weight loss and cachexia. The cause of this hypermetabolism is largely unknown; hypothalamic dysfunction has been discussed in this regard^15,16^. The extent of weight loss is an independent prognostic factor^17,18^ for survival in ALS, i.e., mortality increases by 24% per 1 point loss of BMI^18^. Accordingly, it has been shown that survival of ALS mice can be prolonged by applying a high-caloric nutrition^19^. Furthermore, ALS patients feature distinct alterations of lipid metabolism, and various studies suggest a protective effect of high triglyceride serum levels^20-22^.

Before LIPCAL-ALS-I, evidence for the efficacy of a high-caloric nutrition in human ALS patients has been virtually absent, mainly relying on one single randomized controlled trial in a small number of patients with enteral nutrition via percutaneous endoscopic gastrostomy (PEG)^23^. In LIPCAL-ALS-I, a randomized, placebo-controlled, multicenter trial, beneficial effects of a high-caloric fatty diet (HCFD) for the whole ALS population were demonstrated, including patients in early disease stages^24,25^. Although the primary endpoint (survival in the whole study population) was missed, post-hoc analysis of the primary endpoint, analysis of secondary endpoints, and subgroup analyses revealed very promising and potentially large effects, verifying the proof of concept of this therapeutic approach.

The LIPCAL-ALS I study has shown that

1. HCFD increased survival and reduced weight loss in normal to fast-progressing patients (patients with a functional decline measured by ALS Functional Rating Scale Revised, ALSFRS-R) above the median at baseline; p=0.02); post-hoc analysis showed that these beneficial effects were present in 86% of the whole study population of LIPCAL-ALS I. Hence, excluding only the 14% of slowest progressive patients (corresponding to a progression rate cutoff of 0.25 ALSFRS-R points/month) would have already yielded a significantly positive study result^24^.
2. HCFD slowed down functional decline (measured by ALSFRS-R) in the whole study population during the intervention period; p<0.01^25^.
3. HCFD is the first intervention in sporadic ALS which lowered neurofilament light chain (NfL) serum levels as a prognostic biomarker^26,27^ in the whole study population; p=0.02^25^.

Thus, beneficial effects on all evaluated domains (survival, function, body weight, and biomarkers) were present in the large subgroup of normal to fast-progressing patients. Therefore^24,25^, this study directly aims at the burden of ALS by prolonging survival for the individual patient. Given the scarce treatment options currently available, a positive outcome of LIPCAL-ALS II would yield an immediate and substantial impact on clinical practice, as ultra-high-caloric fatty diet (UFD) would be established as a standard therapy for ALS together with riluzole and constitute one of only two available, disease-modifying treatment options in Europe.

### 1.2 Treatments and rationale for dose selection

#### Treatment Group 1

A dietary supplement consisting of long chain triglyceride fat emulsion (Calogen®) will be administered as per product information. Patients will drink **35 ml four times a day**. At total of 140 ml per day corresponds to an intake of 630 kcal (70 g fat).

Rationale for dose selection: In LIPCAL-ALS I, the intake of 405 kcal and 45 g fat per day in addition to normal food intake was not enough to completely stabilize body weight^24^. However, due to the positive signals obtained in LIPCAL-ALS I, it is hypothesized that survival can be prolonged and that body weight can be completely stabilized by administering the 1.5-fold dosage (630 kcal and 70 g fat = ultra-high-caloric fatty diet, UFD). During the preparational TOLCAL study^28^, a borderline tolerability and drop-out rate (25%) for the 2-fold dosage compared to LIPCAL-ALS I was found. Therefore, only the 1.5-fold amount of calories and fat compared to LIPCAL-ALS I will be applied in LIPCAL-ALS II.

#### Treatment Group 2

A low caloric <5 % fat emulsion (<50 kcal/day) will be used as placebo solution and will be administered in Arm 2. Patients will drink **35 ml four times a day** (140 ml per day).

| Nutritional Facts<br>per 140 ml (Daily Dosage) | Verum (Calogen®) | Placebo |
| --- | --- | --- |
| Energy | 630 kcal | <50 kcal |
| Fat | 100% (70 g) | <5% (<3,5 g) |
| - Saturated fatty acids | 7,5 g |  |
| - Monounsaturated fatty acids | 42,6 g |  |
| - Polyunsaturated fatty acids | 19,9 g |  |
| - Long-chain fatty acids | 100% |  |
| - Ratio omega-6 to<br>omega-3 fatty acids | 5:1 |  |
| protein | 0 g | 0 g |
| carbohydrates | 0 g | 0 g |
| fiber | 0 g | 0 g |

#### Concomitant nutrition

UFD or placebo will be taken in addition to the usual, pre-existing food intake which in ALS implies the recommendation of a high-caloric nutrition. Two distinct standardized scales (CNAQ and Ulm Nutrition questionnaire) will be applied throughout the study to assure that patients maintain their appetite and normal food intake and that patients in both groups do not differ regarding additional food intake.

#### Percutaneous Endoscopic Gastrostomy (PEG)

The nature of the intervention implies an enteral application, either via oral feeding or, alternatively, via PEG. In patients with PEG, analogous to oral feeding, patients will receive the same dosage of 630 kcal in addition to normal food intake. In case of PEG, the term “normal food intake” refers to well-established feeding schemes commonly applied in ALS patients^29^. These feeding schemes generally refer to a high-caloric nutrition aiming at >1500 kcal per day^29^ and therefore mirror the recommendation for high-caloric nutrition in oral-feeding patients.

#### Pre-existing or concomitant high-caloric dietary supplements

If patients already take any dietary supplements before enrollment into this study, the intake of these dietary supplements should be continued during this study. The treating physician may prescribe high-caloric dietary supplements during the study if needed, i.e., with the intention to stabilize body weight. Any treatments (e.g. additional dietary supplements) must be documented in the eCRF.

#### Intake of study intervention

As the study intervention has an unpleasant taste, it is recommended that the study intervention is mixed with a drink, e.g. juice. The study intervention can be taken together with meals or separately. There is no fixed schedule, but patients are advised to maintain their intake times throughout the study to reduce the risk of forgetting a dosage. A reliable scheme is to take 3 dosages during the meals and another dosage in the afternoon, e.g. 8 a.m. (breakfast), 12 p.m. (lunch), 4 p.m. (in between / snack) and 8 p.m. (dinner). There should be a time interval of at least 1 hour between intake of the study intervention and intake of riluzole, as fat-rich nutrition might lower the bioavailability of riluzole.

### 1.3 Summarized risk-benefit assessment

ALS is a devastating disease which dramatically reduces life expectancy. Potential risks of treatments must be assessed against this background. The study intervention (Calogen®) is a commercially available dietary supplement, hence, the overall safety profile in humans is well-known. The precursor study LIPCAL-ALS I proved that Calogen® is safe and well tolerated in ALS^24^. In the preparational TOLCAL trial^28^, the 2-fold dosage compared to LIPCAL-ALS I (3×60 ml) was investigated, and no serious adverse events during the treatment period of 4 weeks were found.

Both placebo and Calogen® can be given as add-on to the standard therapy riluzole, and patients will be instructed to maintain their usual eating habits which in terms of ALS implies a high-caloric nutrition. Therefore, all study patients (intervention and placebo) will receive the best possible standard treatment currently available for ALS and will further benefit from close monitoring of their well-being and course of disease during the regular study visits at the specialized ALS centers. Study evaluations comply with routine, non-invasive assessments for ALS; therefore, the study does not imply additional burden.

In summary, it can be assumed that this study offers a positive benefit to risk ratio for the individual patient. An independent Data Safety Monitoring Board (DSMB) will perform a continuous risk-benefit assessment including detailed analysis of Adverse Reactions and Serious Adverse Events. In case of significant risks, study participation for the individual patient or the entire study will be terminated.

## 2 TRIAL OBJECTIVES

The purpose of this study is to demonstrate that increasing caloric intake and improving nutritional status increases survival of ALS patients

### 2.1 Primary objective

The primary objective of the study is to investigate survival time as a comparison between the control group and experimental group. The survival time will be addressed in terms of the time from randomization until death, tracheostomy or permanent continuous (>22 hours) ventilator dependence.

### 2.2 Secondary objectives

The secondary objectives of the study are to assess the change in disease progression, motor function and quality of life by measuring the change of ALS Functional Rating Scale – Revised (ALSFRS-R) and Rasch Overall ALS Disability Scale (ROADS) from baseline, the effect on Quality of Life (EQ-5D-5L), respiratory function (SVC), body mass index and the Neurofilament Light Chain (NfL) serum concentration, to assess the difference between the observed and predicted ALSFRS-R decrease per month using an NfL-based prediction model^30^, to assess the difference between the observed and predicted survival in NF-ASSESS score groups (Witzel et al., submitted), and to assess the impact of the study intervention on eating habits. Moreover, study contentment and its changes during the study will be evaluated (“Ulmer Fragebogen zur Studienzufriedenheit”).

### 2.3 Assessment of Safety

Assessment of clinical tolerability and safety will be performed by evaluation of terms, frequency, relationship, and seriousness of adverse events (AEs), and serious adverse events (SAEs).

## 3 TRIAL DESIGN

This is a prospective, multicenter, randomized, stratified, parallel-group, double-blind, confirmatory study comparing placebo with an UFD in in 392 enrolled patients with ALS. For entry, the El Escorial criteria for diagnosis of ALS will be used. The patients are required to be on a stable dose of riluzole, or taking no riluzole for the last 4 weeks prior to randomization, to be on a stable dose of edaravone, or taking no edaravone for the last 4 weeks prior to randomization, and to be on a stable dose of sodium-phenylbutyrate/taurursodiol, or taking no sodium-phenylbutyrate/taurursodiol for the last 4 weeks prior to randomization. After patients are screened for inclusion and exclusion criteria, they will be randomly assigned to treatment with either placebo or dietary supplement according to their stratum (progression rate ALSFRS-R ≤0.62 vs. ALSFRS-R ≥0.62 points loss per month; weight loss within 6 months prior to baseline yes/no). Patients will be treated for 18 months in this study. The study requires a combined screening/randomization visit (V1), 4 subsequent control visits (V2-V5), and 5 telephone contacts (T1-T5).

The population consists of 392 enrolled patients with possible, probable, or definite ALS as defined by the El Escorial criteria (revised version)^1^. 196 patients will be randomized to treatment with dietary supplement and 196 patients to treatment with placebo, respectively. The male to female distribution in ALS is 1.1:1. There are no specific requirements concerning the gender distribution within this study as there are no differences in the course of disease. This is a multicenter study including approximately 25 sites in Germany.

### 3.1 Overall trial duration

The estimated duration of the whole study will be about 36 months: 12 months for recruitment, 18 months for the treatment period and 9 months for cleaning the data, carrying out statistical analyses, and writing the final study report. Recruitment of patients will start in Q3 2024, so the anticipated end of study will be in Q3 2027. The actual overall study duration or patient recruitment period may vary from this estimate.

### 3.2 Number of patients

It is planned to enroll **392 patients**, i.e. **196 patients per treatment group**. The sample size calculation is based on a subgroup analysis of the precursor LIPCAL-ALS I study as represented by the proposed inclusion/exclusion criteria (disease progression/ ALSFRS-R slope of ≥0.33 points lost per month at screening and disease duration <36 months). The analysis of this subgroup in LIPCAL-ALS I revealed a survival hazard ratio of 0.61 for HCFD (405 kcal) vs. placebo after 18 months. The effect for UFD (630 kcal) vs. placebo is unknown, but, due to the increased dosage, at least the same effect as seen for HCFD in LIPCAL-ALS I can be assumed, corresponding to a survival rate of 47% in the placebo group and 63% in the treatment group after 18 months. With an assumed follow-up time of 18 months for each patient (no accrual), a two-sided significance level of 5%, and a power of 80%, a sample size of 290 patients (145 per treatment group) will have to be analyzed. 130 events will be needed. Assuming a dropout rate of 26% as seen in LIPCAL-ALS I^24^, 392 patients (196 per group) will have to be randomized.

Due to the otherwise broad inclusion/exclusion criteria and based on previous trials^31,32^, 5% of additional screening failures are expected, corresponding to a total sample size of 412 patients to be assessed for eligibility.

### 3.3 Primary endpoint

The primary endpoint is survival (time to death, tracheostomy or permanent continuous (>22 hours) ventilator dependence).

### 3.4 Secondary endpoints

- Change of total score of ALSFRS-R^2^
- Change of linearly-weighted normed ROADS score^3^
- Change of Quality of Life (EQ-5D-5L)
- Change of SVC
- Survival (time to death, tracheostomy or permanent continuous (>22 hours) ventilator dependence) after 6 and 12 months
- Time to death
- Time to tracheostomy
- Time to permanent continuous ventilator dependence (>22 hours)
- Ventilation assistance-free survival (VAFS)
- Change of BMI
- Change of appetite (CNAQ)
- Change of eating habits (UNQ)
- Change of NfL serum concentration
- Difference between the observed and predicted ALFRS-R decrease per month; predicted ALSFRS-R decrease will be computed using a validated serum NfL-based prediction model^30^
- Difference between the observed and predicted survival in NF-ASSESS score groups (Witzel et al., submitted)

### 3.5 Exploratory endpoints

- The following biomarkers constitute promising, but not established potential biomarkers for neurodegenerative or inflammatory processes of neurons, synapses, and glia cells, and will be measured in serum and CSF: MAP2, UCHL1, GPNMB, YKL-40, SNAP 25, beta-synuclein, Aquaporin-4, GFAP, sTREM, CCL2, IL1b, IL2, IL4, IL6, IL10, IL12p70, IL17, TNF-alpha.
- Study contentment (“Ulmer Fragebogen zur Studienzufriedenheit) and its change during the study

### 3.6 Safety variables

Safety and tolerability of the dietary supplement will be investigated by analyzing terms and frequencies of AEs and SAEs.

### 3.7 Measures taken to minimize/avoid bias

#### 3.7.1 Randomization and Stratification

This study has two parallel groups (dietary supplement versus placebo), is randomized, and treatment groups are completely masked (double-blind study). The randomization will be performed stratified according to ALSFRS-R progression rate pre-baseline (≤**0.62 vs**. ≥**0.62 points lost per month**) and weight loss (**yes/no during 6 months prior to baseline**), because these parameters are suspected to affect treatment efficacy. For the assessment of stratification factors, the patients’ medical data or (if not available) the patients’ recollection will be used. The ALSFRS-R progression rate will be calculated by the formula: **[48 – (ALSFRS-R at screening visit)] / months between disease onset and screening visit**. The disease onset is defined as the occurrence of first paresis / muscle weakness (in bulbar patients: first occurrence of speech or swallowing difficulties).

The randomization list will be generated by the Interdisciplinary Centre Clinical Trials (IZKS) Mainz, Germany, using a validated system, which involves a pseudo-random number generator to ensure that the resulting treatment sequence will be both reproducible and non-predictable^33^. Dietary supplement (Calogen®) is commercially available and will be blinded by the Pharmacy of the University Hospital Ulm, Germany. Calogen® and placebo solution will be manufactured by Nutricia GmbH, Erlangen, Germany. All blinded dietary supplements will be sent directly to the patients. The randomization list will be kept in safe and confidential custody at Pharmacy of the University Hospital Ulm, Germany.

#### 3.7.2 Blinding

Blinding of both the investigator and patient is achieved by providing dietary supplement and matching placebo solution. Both Calogen® and placebo are white milky solutions with similar taste.

The investigator will receive an emergency number to reach the pharmacist on duty. The pharmacist on duty will be able to provide information on the patient’s study intervention. Date and reason for unblinding must be documented by the investigator or an authorized person in the eCRF and on the medical record of the patient.

### 3.8 Selection and withdrawal of patients

No patient will be allowed to enroll in this study more than once.

#### 3.8.1 Inclusion criteria

- Possible, probable (clinically or laboratory supported) or definite ALS according to the revised version of the El Escorial criteria^1^
- Disease duration (onset of first paresis or bulbar symptoms) < 36 months
- Loss of ALS functional rating scale revised (ALSFRS-R) of ≥ 0.33 points/month based on the formula:
- (48 – ALSFRS-R at Screening Visit) / (Months between Onset and Screening Visit)
- Age ≥18 years.
- Either continuously treated with a stable dose of riluzole, OR not treated with riluzole for the last 4 weeks prior to inclusion
- Either continuously treated with a stable dose of edaravone, OR not treated with edaravone for the last 4 weeks prior to inclusion
- Either continuously treated with a stable dose of sodium-phenylbutyrate/taurursodiol, OR not treated with sodium-phenylbutyrate/taurursodiol for the last 4 weeks prior to inclusion
- Capable of thoroughly understanding all information given
- full informed consent according to good clinical practice (GCP).

#### 3.8.2 Exclusion criteria

Patients presenting with any of the following criteria will not be included in the study:

- Previous participation in another interventional study involving an active treatment within the preceding 4 weeks
- Tracheostomy or continuous permanent ventilator dependence (>22 hours per day)
- Pregnancy or breastfeeding
- Any medical condition known to have an association with motor neuron dysfunction which might confound or obscure the diagnosis of ALS
- Presence of any concomitant life-threatening disease or impairment likely to interfere with functional assessment.
- Evidence of a major psychiatric disorder or clinically evident dementia precluding evaluation of symptoms.
- Liable to be not cooperative or comply with study requirements as assessed by the investigator, or unable to be reached in the case of emergency

#### 3.8.3 Withdrawal criteria

Patients may withdraw their consent at their own request without giving reasons during the study without any disadvantages. Whenever possible, all patients prematurely discontinuing study participation should perform an early termination visit including the same evaluations as scheduled for visit V5. The reason(s) for premature cessation of a patient’s participation during the study should be specified on the case report form. Patients terminating the study within the first 2 weeks will be replaced. If a patient is no longer able to visit the study center due to progressing disease or any other reasons, the primary endpoint survival will still be recorded.

In addition, patients will be discontinued from the study under the following circumstances:

- enrollment in any other clinical trial involving an investigational product or enrollment in any other type of medical research judged not to be scientifically or medically compatible with this study.
- on Investigator’s decision in case of any clinical adverse event (AE), laboratory abnormality or intercurrent illness, which, in the opinion of the investigator, indicates that continued participation in the study is not in the best interest of the patient.
- on Sponsor’s decision if Sponsor stops the whole study, in case of premature closure of trial site, if the patient’s participation in the study cannot be continued for medical, safety, regulatory, or other reasons consistent with applicable laws and regulations, or if the patient is non-compliant with study procedures and/or treatment

For any patient prematurely discontinuing the study participation, the investigator must:

- As far as possible, carry out all the examinations and tests planned for the final visit after withdrawal
- Fill in the corresponding pages of the case report form, specifying the date and reason for discontinuation
- When applicable, ask the patient for the reason of his/her informed consent withdrawal while fully respecting the patient’s rights
- When possible, conduct an on-site visit according to V5. If an on-site visit cannot be performed, conduct a phone visit and collect all outcome parameters collectable via phone.
- All ongoing serious adverse events of withdrawn patients must be followed up until no more signs and symptoms are verifiable or the investigator has assessed its sequelae, even after the end of the study.

#### 3.8.4 Premature closure of study sites

For the following reasons a study site may be closed at the discretion of the sponsor:

- Medical or ethical reasons which might have disadvantageous effect for the whole study
- Difficulties in the recruitment of subjects
- Critical protocol violations
- Violations of legal and ethical regulations
- Non-compliance of the study site investigators

#### 3.8.5 Premature closure of the clinical study

For the following reasons the whole study may be discontinued at the discretion of the sponsor:

- New risks for patients become known.
- Inefficacy of the study intervention becomes evident.
- Occurrence of AEs unknown to date in respect of their nature, severity, and duration or the unexpected increase in the incidence of known AEs.
- Medical or ethical reasons affecting disadvantageously the continued performance of the study.
- Difficulties in the recruitment of patients.

The ethic committees must then be informed. Should the study be closed prematurely, all study material must be returned to the sponsor or will be collected by the monitor.

## 4 TRIAL TREATMENTS

### 4.1 Treatment assignment

Following completion of screening assessments and randomization, four times daily oral administration of dietary supplement will be initiated for each patient, consisting of UFD or placebo. Patients withdrawn from the study retain their identification codes (e.g. randomization number). New patients will always receive a new identification code.

### 4.2 Dosage schedule

The recommended dosage (maximum dosage) of study treatment is 140 ml/day (35 ml four times a day). Dose adjustments, e.g. dose reduction in case of side effects, are allowed. In case of persisting tolerability issues, it is recommended to reduce the dosage to 3×35 ml. If tolerability issues persist after dosage reduction, further measures should be discussed with the sponsor. A flexible dose regimen should balance treatment adherence and feasibility with the conduct of protocol treatment. The exact dosage must be documented in the CRF.

### 4.3 Procedures for monitoring patient compliance

The UNQ will be filled out to determine the patient’s compliance.

## 5 TRIAL SCHEDULE

### 5.1 Trial Flow

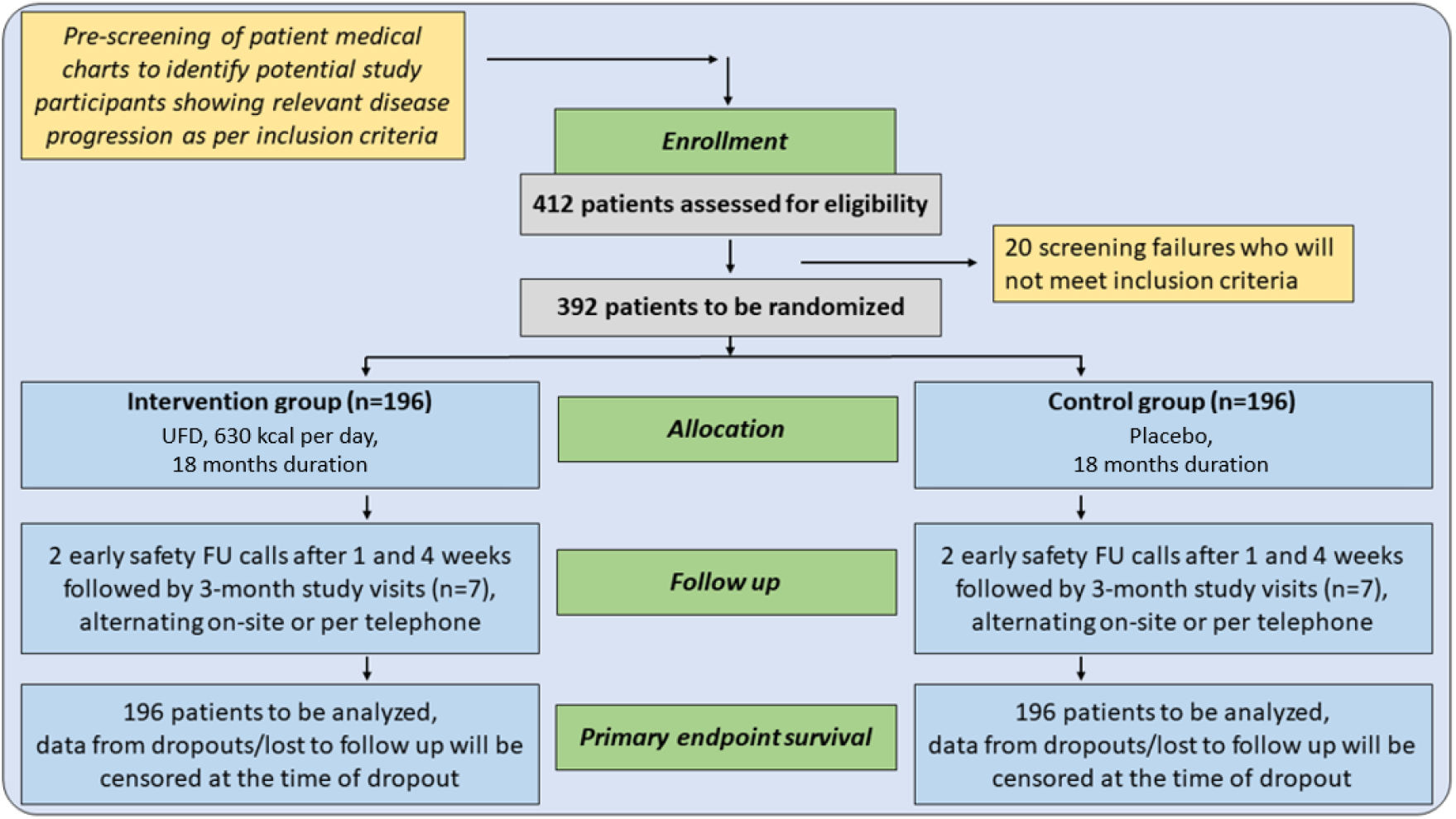

### 5.2 Trial visits schedule

There is no distinct baseline visit, i.e., screening and baseline visit will be conducted as one visit. For each visit, the date of the screening visit (enrollment in the study) is considered as the reference date. The dates of the following visits specified in the protocol are calculated in relation to the date of this visit. If the time of a visit is modified in relation to the calculated schedule, the times of the following visits will always be determined with respect to the screening visit. The time window for each planned visit will be **± 2 weeks** except for T1 and T2, which should be performed within ± 2 days). The visits schedule is not modified by any unscheduled visits. If a patient misses one onsite or phone visit, the subsequent visit will be carried out as scheduled. If a patient misses more than one onsite or phone visit, the investigator should contact the sponsor to determine how to proceed.

#### Screening visit (V1)

A signed “Informed Consent” will be obtained from each patient prior to initiating any study procedure. Presence of ALS diagnostic criteria will be documented as well as site of onset and ALS status at entry according to the revised El Escorial criteria. Venous blood will be taken for the determination of blood count, electrolytes, liver and kidney values, CRP, glucose, Hba1c, cholesterol, HDL, LDL, triglycerides, CK, NfL, and exploratory/scientific parameters. A pregnancy test (rapid test in urine is sufficient) will be conducted. Demographics and a detailed medical history will be taken, and concomitant medication will be recorded. A physical examination, including body weight, height, BMI, blood pressure, and radial pulse will be performed. Additionally, lung function will be assessed by SVC. Additionally, lung function will be assessed by SVC. In addition, ALSFRS-R, ROADS score, EQ-5D-5L, CNAQ, UNQ and “Ulmer Fragebogen zur Studienzufriedenheit” will be performed. The patient’s eligibility will be determined in accordance with the inclusion/exclusion criteria. A lumbar puncture can be conducted on a voluntary basis to obtain CSF for exploratory/scientific purposes.

Patients eligible for study participation will be randomized to one of the treatment groups. Acquired data will be utilized as baseline measurements.

#### Onsite Visits (V2–V5)

Visits 2–5 will be performed 3, 6, 12, and 18 months after V1, respectively. Venous blood samples will be taken (see Screening Visit). At V3 and V4, a lumbar puncture can be conducted on a voluntary basis to obtain CSF for exploratory/scientific purposes. A physical examination (including vital signs, height, weight, blood pressure, heart rate, internal and neurological examination) will be conducted, and lung function will be assessed by SVC. In addition, ALSFRS-R, ROADS score, EQ-5D-5L, CNAQ, UNQ, and “Ulmer Fragebogen zur Studienzufriedenheit” will be performed. Any changes in the concomitant medication and adverse events will be documented, and compliance to the study protocol will be checked.

#### Phone Visits (T1 – T5)

After 5 days, after 2 weeks, and after 1, 9, and 15 months, the patients or (in case of severe dysarthria / anarthria) their family members will be contacted by phone to assess ALSFRS-R (T4+T5), ROADS score (T4+T5), CNAQ (T2–T5), UNQ (T2–T5), adverse events (T1–T5), changes in concomitant medication (T2–T5), and compliance to the study protocol (T1–T5).

## 6 TRIAL METHODS

### 6.1 Assessment of efficacy

Efficacy variables:

- Survival (date of death)
- ALSFRS-R
- ROADS score
- EQ-5D-5L
- SVC
- Date of Tracheostomy
- Date of permanent continuous (>22 hours) ventilator dependence
- BMI
- CNAQ
- UNQ
- Ulmer Fragebogen zur Studienzufriedenheit
- NfL serum concentration
- Difference between the observed and predicted ALSFRS-R decrease per month; predicted ALSFRS-R decrease will be computed using a validated serum NfL-based prediction model^30^
- Difference between the observed and predicted survival in NF-ASSESS score groups (Witzel et al., submitted)

### 6.2 Assessment of safety

#### 6.2.1 Safety Variables

- Blood cell count (RBC, WBC, Hb, Hct)
- Electrolytes
- Liver Values (AST, ALT, GGT)
- Creatinine
- CRP
- Glucose, Hba1c
- Cholesterol, HDL, LDL, Triglycerides
- CK

#### 6.2.2 Adverse events

##### 6.2.2.1 Definitions

###### Adverse Event (AE)

According to GCP, an adverse event (AE) is defined as any untoward medical occurrence in a patient treated with an investigational product and which does not necessarily have a causal relationship with this treatment. An AE can therefore be any unfavorable and unintended sign (including an abnormal laboratory finding), symptom, or disease, temporarily associated with the use of an investigational product, whether related to that product.

An AE may be:

- a new symptom or medical condition
- a new diagnosis
- a change in laboratory parameters
- an intercurrent illness or accident
- worsening of a medical condition/disease existing before the start of the clinical study
- recurrence of a disease
- an increase in frequency or intensity of episodic diseases.

This definition applies to events occurring in any comparative group (active treatment or placebo), including periods when the patient is not receiving any study product.

Surgical procedures themselves are not AEs; they are therapeutic measures for conditions that require surgery. The condition for which the surgery is required may be an AE. The condition(s) leading to these measures are not AEs, if the condition leading to the measure was present before inclusion in the study. In the latter case the condition should be reported as medical history.

The criteria for determining whether an abnormal laboratory test finding should be reported as an adverse event are as follows:

- Test result is associated with accompanying symptoms, and/or
- Test result requires additional diagnostic testing or medical/surgical intervention, and/or
- Test result leads to a change in study dosing outside of protocol-stipulated dose adjustments, or discontinuation from the study, significant additional concomitant drug treatment, or other therapy, and/or
- Test result is an adverse event by the investigator or sponsor.

###### Serious adverse event (SAE)

The terms “serious” and “severe” are not synonymous but are often used interchangeably. The term “severe” is often used to describe the intensity (severity) of a specific event; the event itself, however, may be of relatively minor significance (such as severe headache). This is not the same as “serious”, which is based on patient/event outcome or action criteria usually associated with events that pose a threat to a patient’s life or functioning. Seriousness (not severity) serves as a guide for defining regulatory reporting obligations.

Due to the natural disease progression of ALS, hospitalizations occur more frequently than in most other indications. For example, throughout the course of the disease, patients may suffer from dysphagia or respiratory insufficiency, which may result in hospitalizations for the application of PEG or NIV, respectively. Such events shall be documented as such in the patient’s CRF; however, they are **not** considered as an SAE in this study. On the other hand, events which are related to ALS, but are not obligatorily associated with natural disease progression (e.g., aspiration pneumonia, fall) **are** considered SAEs, if they fulfill any of the criteria below.

A serious adverse event (SAE) is an adverse event that fulfills any of the following criteria:

- results in **death**
- is **life-threatening**, i.e., the patient was at immediate risk of death at the time of the serious adverse event; it does not refer to a serious adverse event that hypothetically might have caused death if it had been more severe
- requires patient **hospitalization** or prolongation of existing hospitalization; in any of the following situations, the serious criterion for hospitalization is <u>not</u> fulfilled:
  - the admission was pre-planned (i.e., elective, or scheduled surgery arranged prior to start of the study)
  - the admission was not associated with an adverse event (e.g., hospitalization for social purpose)
  - the stay in hospital was for less than 12 hours; however, in this case, the criteria for other SAE criteria might still be fulfilled (e.g. “medically important”, see below)
- results in persistent or significant **disability/incapacity**, i.e., there is a substantial disruption of a person’s ability to carry out normal life functions. The irreversible injury of an organ function (e.g., paresis, diabetes, cardiac arrhythmia) fulfills this criterion.
- study participant / partner of study participant gives birth to a child with **congenital anomaly/birth defect**
- is an **important medical event**. Deciding whether expedited reporting is appropriate in situations where none of the outcomes listed above occurred should be based on medical and scientific judgement. Important medical events that may not be immediately life-threatening and do not result in death or hospitalization, but may jeopardize the patient’s health or may require intervention to prevent one of the other outcomes listed in the definition above should also be considered serious. Examples of such events include allergic bronchospasm requiring intensive treatment in an emergency room or at home, diagnosis of cancer during the treatment, or the development of drug dependency or drug abuse.

##### 6.2.2.2 Assessment of Adverse Events

Patients must be carefully monitored for adverse events by the investigator. The intensity of adverse events and the causal relation to the intervention must be assessed.

###### Intensity/Severity

The intensity of an AE will be assessed by the investigator as follows:

- <u>Mild</u>: Temporary well tolerated by the patient and does not interfere with normal daily activities
- <u>Moderate</u>: results in discomfort for the patient and impairs his/her normal activity
- <u>Severe:</u> results in substantial impairment of normal activities

###### Causal relation to study procedures/dietary supplement

The assessment of the relationship between an AE (clinical event or laboratory test) and the intervention requires a clinical judgement based on all available information at the time of the completion of the CRF according to modified criteria of WHO from 1991:

- <u>Certain</u>: occurred in a plausible time relationship to the intervention and cannot be explained by concurrent disease or other drugs or chemicals.
- <u>Probable:</u> occurred with a plausible time relationship to the intervention, but could also be explained by concurrent disease or concomitant drugs or chemicals.
- <u>Possible:</u> does not fulfill the criteria of any other category
- <u>Unlikely:</u> the temporal relationship to the intervention makes a causal relationship improbable, and other drugs, chemicals or underlying disease provide plausible causes
- <u>Not related:</u> caused by the patient’s clinical state, other modes of therapy, or any other known etiology

###### Outcome

The outcome of an AE must be assessed according to the following classification:

- <u>Completely recovered</u>: recovered with no observable residual effects
- <u>Not yet completely recovered</u>: improved, but still some residual effects
- <u>Permanent damage:</u> suspected permanent impairment
- <u>Death</u>
- <u>Ongoing:</u> AE has not been resolved
- <u>Unknown:</u> lost to follow-up

##### 6.2.2.3 Observation Period

The observation period for collection of adverse events begins with the signature of the informed consent and **ends 14 days after the patient has terminated the study**, either routinely or prematurely. If the investigator becomes aware of an SAE after the observation period and considers the event as possibly, probably, or certainly related to the prior study (see above), the investigator should contact the sponsor to determine how the adverse event should be documented and reported.

##### 6.2.2.4 Documentation of Adverse Events and Follow-Up

At each visit after V1, the investigator must question the patient about the occurrence of any adverse event and note any abnormal laboratory findings. For this purpose, a neutral question such as “Since the last visit, have you felt any untoward or unusual symptoms other than those related to your illness?” should be asked.

All AEs reported by the patient or detected by the investigator must be documented on the appropriate pages of the CRF. AEs must also be documented in the patient’s medical records.

Every attempt should be made to describe the AE in terms of a clearly defined diagnosis. If a clearly defined diagnosis was made, individual signs and symptoms must not be recorded, unless they represent atypical or extreme manifestations, in which case they should be reported as separate AEs. If a clearly defined diagnosis cannot be established, each sign and symptom must be recorded individually.

All patients who have AEs, whether considered as associated with the intervention or not, must be monitored to determine the outcome. The clinical course of the AE must be followed up according to accepted standards of medical practice, even after the end of the observation period, until a satisfactory explanation is found or the investigator considers it medically justifiable to terminate follow-up. Should the adverse event result in death, the pathologist’s report should be supplied, if possible.

##### 6.2.2.5 SAE Reporting

SAEs must be documented in the eCRF. The SAE report should be as complete as possible including the patient’s identification number, the SAE (medical term, diagnosis), and the assessment regarding a causal relationship between the event and the intervention. The SAE report must be reviewed and signed by the investigator.

##### 6.2.2.6 Safety evaluation and reporting by sponsor

The sponsor is responsible for the continuous safety evaluation of the intervention and the clinical study. During the study, the *DSMB* will review the safety data every 6 months and provide a recommendation to the sponsor whether to continue, modify, or terminate the study.

##### 6.2.2.7 Emergency procedures and unblinding

During and after a patient’s participation in the study, the investigator should ensure that adequate medical care is provided for any AEs, including clinically significant laboratory values. The investigator must inform a patient whether medical care is needed for any AE of which the investigator becomes aware.

In case of an emergency, the investigator will receive information about the study intervention (UFD or placebo) by the pharmacist of duty of Ulm University Clinic. Contact details of the pharmacist of duty and the process flow for unblinding will be handed out to each study site during the site initiation visit. No other reason than an emergency may justify unblinding. After unblinding, the investigator must note the date, time, and reason for unblinding in the CRF. The investigator must also immediately inform the study monitor. Whenever possible, the coordinating investigator, Prof. Dr. Johannes Dorst, should be contacted before the blind is broken.

### 6.3 Other assessments

#### 6.3.1 Prior and concomitant diseases

Relevant other diseases present at the time of informed consent are regarded as concomitant illnesses and will be documented on the appropriate pages of the CRF, as well as clinically significant prior conditions within the last 10 years.

#### 6.3.2 Prior and concomitant treatments

Relevant additional treatments administered to the patients at the time of screening or at any time during the study are regarded as concomitant treatments und must be documented on the appropriate pages of the CRF.

#### 6.3.3 Blood sampling for scientific testing

During the scheduled on-site visits blood will be drawn for the assessment of several safety measures and for NfL concentrations. Additionally, at each visit, an amount of approximately 40 ml blood (30 ml serum and 10 ml EDTA-plasma) will be drawn from patients for further exploratory scientific analysis. At each visit, patients may also agree to undergo a lumbar puncture to provide CSF samples. This measure is entirely voluntary, i.e., patients who do not agree to this measure may still participate in the study.

These samples will be utilized to address a variety of scientific questions, especially the effect of UHD on various metabolic and neurodegenerative biomarkers, with the goal to facilitate earlier diagnosis, improved disease monitoring, and establishing additional suitable outcome parameters for clinical studies.

#### 6.3.4 Compliance

Based on the experience from previous trials^24,28^, compliance / adherence to study treatment is an important issue in clinical studies with nutritional interventions. Nutritional interventions may have an unfavorable taste and cause gastrointestinal side effects. Moreover, patients tend to underestimate the potential power of a nutritional intervention. These factors should already be discussed with the patient during the initial screening visit as follows:

- the potential power of a nutritional intervention should be explained (including the promising results from LIPCAL-ALS I)
- it should be explained that the nutritional intervention should be considered as treatment (equivalent to a drug) rather than food
- it should be mentioned that, although the patient may terminate the study at any point of time, the patient should only participate if he/she is confident that he/she is able to complete the study
- it should be mentioned that mild tolerability issues are frequent, but commonly disappear after a few days/weeks.
- It should be explained that the intervention may have an unfavorable taste if consumed purely, but that the intervention may be mixed with any drink, such as juice, milk, tea, or coffee
- Patients should be instructed that in case of problems they shall not terminate the study by themselves, but instead contact the responsible physician or study nurse to discuss possible solutions.

Moreover, two early phone visits after 5 days and 2 weeks (T1 and T2) have been implemented. During these phone visits, the conducting investigator or study nurse should thoroughly evaluate tolerability issues and compliance, and should provide advice if necessary, including:

- In case of complaints about the <u>taste</u>: inform about the possibility to mix with drinks (see above); explain the importance and potential power of a nutritional intervention
- In case of <u>mild tolerability issues</u>: explain that mild gastrointestinal symptoms tend to disappear after a few days
- In case of <u>significant tolerability issues</u>: consider to reduce the dosage from 4×35 ml to 3×35 ml; in case the tolerability issues persist despite dosage reduction, the investigator/study nurse should discuss the case with the sponsor to evaluate further adjustments (e.g. further dosage reduction, change of dosage distribution, change of intake times)

If the investigator or study nurse becomes aware of any tolerability or compliance issues during the phone or on-site visits, he/she should follow-up on these issues, i.e., arrange follow-up phone calls with the patients and evaluate whether the problem has been resolved by the measures given above. In any case of doubt, investigators are encouraged to contact the sponsor how to proceed. Any compliance or tolerability issues must be documented in the CRF.

## 7 STATISTICS

Details of the statistical analysis of the data collected in this study will be documented in the SAP that will be provided by the study statistician and finalized before closing the data base and prior to breaking the blind. The SAP is based on the protocol including all amendments. The document may modify the plans outlined in this protocol; however, any major modifications of the primary endpoint definition and/or its analysis will also be reflected in a protocol amendment. Any deviation from the original statistical plan must be described and justified in the final report. The statistical analysis will be conducted using SAS^®^.

### 7.1 Sample size

The sample size calculation is based on a subgroup analysis of the precursor LIPCAL-ALS I study^24^ represented by the proposed inclusion/exclusion criteria (disease progression/ ALSFRS-R slope of ≥0.33 points lost per month at screening and disease duration <36 months). The analysis of this subgroup revealed a survival hazard ratio of 0.61 for HCFD (405 kcal) vs. placebo after 18 months^24^. The effect for UFD (630 kcal) vs. placebo treatment is unknown, but due to the increased dosage, at least the same effect as seen for HCFD in LIPCAL-ALS I can be assumed, corresponding to a survival rate of 47% in the placebo group and 63% in the treatment group after 18 months^24^. With an assumed follow-up time of 18 months for each patient (no accrual), a two-sided significance level of 5%, and a power of 80%, a sample size of 290 patients (145 per treatment group) will have to be analyzed. 130 events will be needed. Assuming a dropout rate of 26% as seen in LIPCAL-ALS I^24^, 392 patients (196 per group) will have to be randomized. Due to the otherwise broad inclusion/exclusion criteria and based on previous trials^31,32^, 5% of additional screening failures are expected, corresponding to a total sample size of 412 patients to be assessed for eligibility.

### 7.2 Analysis populations

All patients who signed informed consent and received a randomization number are considered as enrolled, even if they did not receive any study treatment. The following data sets will be analyzed:

- <u>Full Analysis Set (FAS)</u>: The primary efficacy analysis will be based on the Intention-To-Treat (ITT) population. All randomized patients who received at least one dose of study treatment and with at least one available post-baseline assessment of the primary analysis variable will be included in the FAS and will be regarded as ITT population. This population is the primary analysis population. Within the ITT population analysis, patients will be assigned to the treatment to which they were randomized.
- <u>Per-protocol Analysis Set (PPS)</u>: The PPS will consist of all patients in the full analysis set without major protocol deviations. Patients with major protocol deviations will be excluded from the PPS. Specifically, patients in the PPS are required to have taken >65% of the intended total dietary supplement volume throughout the study and to fulfill all inclusion and exclusion criteria. A more detailed description of protocol deviations and analysis sets will be included in the statistical analysis plan which will be finalized before unblinding the treatment allocation and before start of the statistical analyzes. The final definition of minor and major protocol deviations and the resulting definition of analysis sets will be provided and approved by the sponsor prior to final statistical analysis and prior to unblinding of the study. A per-protocol analysis will be performed for the primary endpoint, but the primary analysis set for the efficacy evaluation is the FAS. The per protocol analysis will be considered as supportive since it might violate the comparability of the groups achieved by randomization.
- <u>Safety Analysis Set</u>: The safety analysis set comprises all patients who received at least one dose of study treatment. In the Safety Analysis Set, patients will be assigned to the treatment which they received.

### 7.3 Statistical Analysis

The primary objective of this study is to show the superiority of the test treatment compared to the standard treatment in terms of survival time.

#### 7.3.1 Definition and analysis of primary endpoint

For primary analyses, a Cox regression analysis with the factors treatment group, ALSFRS-R slope before baseline (≤0.62 points lost per month versus ≥ 0.62 points lost per month, corresponding to the median of LIPCAL-ALS I^24^), NfL serum levels at baseline, and weight loss within 6 months prior to baseline (yes/no) will be used. Models will be fitted for experimental group versus placebo. The model assumptions are justified according to the precursor trial LIPCAL-ALS I. Nevertheless, Schoenfeld residuals will be checked. For robustness, Cox regression models with additional covariates (e.g. center and sex) will be analyzed.

Treatment differences will be displayed by hazard ratios and their two-sided 95% confidence intervals. If the hazards are non-proportional, a restricted mean survival time approach will be applied. Kaplan-Meier survival curves will be presented. Each comparison will be conducted on a two-sided level of significance of 5%. Analyses will be based on all randomized patients (intention-to-treat). In general, patients who discontinued the trial before an event are considered censored. However, patients dropping out prematurely for massive disease progression will be checked for informative censoring. These patients might be considered for experiencing an event. Losses to drop-out will be handled as censored at the time point of last contact. The analysis model uses the data until drop-out. Patients who are non-compliant will be included in the ITT population, but excluded in a per protocol analysis (per protocol set: compliance to dietary supplement more than 65% and adherence to the inclusion/exclusion criteria). The per protocol analysis will be considered as supportive since it might violate the comparability of the groups achieved by randomization.

#### 7.3.2 Analysis of secondary endpoints

Since the primary endpoint is composite, the primary analysis will be repeated for each component (time to death, time to tracheostomy, time to permanent continuous (>22h) ventilator dependence). All secondary parameters will be evaluated by mixed models with repeated measure (MMRM). MMRMs account for missing values by using the correlation of measurements of the observed values. Compound symmetry or an AR (1) covariance structure will be assumed. Descriptive statistics will be provided.

#### 7.3.3 Analysis of Subgroups

For subgroup analysis, a Cox regression model will be fitted with an additional term for the subgroup and the interaction term of treatment group and subgroup in the model. There are subgroup analyses planned based on weight loss within 6 months pre-baseline (yes/no), progression rate pre-baseline (ALSFRS-R loss per month with median and upper third as cut-offs), sex (male and female), and patients with/without PEG pre-baseline. Additional subgroups might be defined in a statistical analysis plan (SAP).

#### 7.3.4 Safety Analyses

Adverse Events will be coded by MedDRA terminology and analyzed by evaluating absolute and relative frequencies. Vital signs and laboratory values will be analyzed by t-tests and sample characteristics. Analysis of further safety parameters

#### 7.3.5 Analysis of clinical laboratory findings

Listings will be prepared for each laboratory measure and will be structured to permit review of the data per patient as they progress on treatment. Summary tables will be prepared to examine the changes of laboratory measures over time. Additionally, shift tables will be provided to examine the changes of laboratory data from normal baseline to values outside the corresponding reference range during/after treatment.

#### 7.3.6 Interim analyses

No interim analyses are planned.

## 8 QUALITY CONTROL AND QUALITY ASSURANCE

### 8.1 Requirements for investigational sites and staff

The investigator should be able to demonstrate (e.g. based on retrospective data) a potential for recruiting the required number of suitable patients within the agreed recruitment period. The investigator should have sufficient time to properly conduct and complete the study within the agreed study period.

The investigator should have available an adequate number of qualified staff and adequate facilities for the foreseen duration of the study to conduct the study properly and safely. The investigator should ensure that all persons assisting with the study are adequately qualified, informed about the protocol, any amendments to the protocol, the study treatments, and their study-related duties and functions.

### 8.2 Direct entries

Data entries to be entered in the CRF as Direct entries are listed in the Monitor Manual in the section source data control.

### 8.3 Direct access to source data/documents

The investigator/institution must permit study-related monitoring and auditing by the sponsor and the IZKS Mainz, as well as inspections by the appropriate authorities, providing direct access to source data/documents.

The patients will be informed that representatives of the sponsor or appropriate authorities may inspect their medical records to verify the information collected, and that all personal information made available for inspection will be handled in strictest confidence and in accordance with the Datenschutz-Grundverordnung (DSGVO).

### 8.4 Investigator site file and archiving

The investigator will be provided with an investigator site file (ISF) at the start of the study. The investigator will archive all study data and relevant correspondence in the ISF. The ISF, all source data and all documents will be kept filed according to the requirements of the ICH-GCP guidelines after termination of the study.

It is the responsibility of the investigator to ensure that the patient-identification sheets are stored for at least 15 years beyond the end of the research project. All original patient files must be stored for the longest possible time permitted by the regulations at the hospital, research institute, or practice in question. If archiving can no longer be maintained at the site, the investigator must notify the sponsor.

### 8.5 Monitoring

Monitoring will be done by personal visits from a clinical monitor according to SOPs of the IZKS Mainz. To initiate the study, the monitor will visit all participating study sites. The monitor must ensure that the investigators and their staff understand all requirements of the protocol and their regulatory responsibilities. Each site will be visited by the monitor at regular intervals to ensure compliance with the study protocol, GCP and legal aspects. After all patients have finished treatment/follow-up, the study is terminated and a close-out visit will be performed.

The monitor will review the entries into the CRFs for completeness and correctness and verify the entries based on the source documents. The presence of correct informed consents will be checked for every patient. Details will be specified in the monitoring manual for this study.

The investigator must allow the monitor to investigate all relevant documents and must always provide support to the monitor. By frequent communications (letters, phone, fax), the monitor will ensure that the study is conducted according to the protocol and regulatory requirements.

## 9 DATA MANAGEMENT

### 9.1 Responsibilities

In case of discrepancies or correction of data errors, the data management team is authorized to contact the responsible person at the study site directly. The queries will be sent by the IZKS Mainz per fax or e-mail. For the response, the investigator must comply with the terms defined by the IZKS Mainz. The investigator must agree to be contacted by e-mail or phone.

A detailed methodology for the data management in this study will be documented in a data management plan (DMP) that will be dated and maintained by IZKS Mainz. This plan must be signed by the sponsor, the head of the data management team and the responsible data manager. The document may modify the plans outlined in this protocol; however, any major modifications of the data handling will also be reflected in a protocol amendment.

### 9.2 Data collection

This study will be performed using an electronic case report form (eCRF) or remote data entry (RDE). The investigator and the study site staff will receive system documentation, training, and support for the use of the eCRF. In case of new study site staff, the training can be performed by personnel of the study site. For support with data entry, the IZKS Mainz can be contacted between 9:00 and 16:00 Monday to Thursday and from 9:00 to 12:00 on Fridays. Each study site has one responsible person who supports the IZKS Mainz regarding the implementation of technical and organizational processes.

All protocol-required information collected during the study must be entered by the investigator or a designated representative in the eCRF. All data entry, modification, or deletion will be recorded automatically in an electronic audit trail indicating the individual patient, the original value and the new value, the reason for change, who made the change, and time and date of the change. All data changes will be clearly indicated. Former values can be viewed in the audit trail. All electronic data will be entered by the site (including an electronic audit trail) in compliance with applicable record retention regulations.

The system will be secured to prevent unauthorized access to the data or the system. Only people provided with a user ID and a password will be able to enter or change data. The investigator will maintain a list of individuals who are authorized to enter or correct data and their system ID.

Computer hardware and software (for accessing the data) will be maintained at or made available for the site in compliance with applicable regulations. All technical preconditions for each study site are recorded in the DMP.

The system can produce exact copies of data in legible paper form for inspections and audits. The principal investigator or a designated investigator, following review of the data in the eCRF, will confirm the validity of each patient’s data by electronic signature or by signing a paper printout of a listing of all patients enrolled in the study. The architecture of the computer system will be described in the DMP.

### 9.3 Data handling

During data entry, integrity checks will help to minimize entry failures. These data entry checks are based on the data validation plan (DVP), signed by the coordinating investigator. The data entry system allows the study monitors to control the entry process with the help of the built-in review functions. Comments and requests can be promptly processed by the study site. Checks for plausibility, consistency and completeness of the data will be performed during data entry. Based on these checks, queries will be produced. Any missing data or inconsistencies will be reported back to the respective site and clarified by the responsible investigator.

After completion of data entry and if no further corrections are to be made in the database, the access rights will expire, and the database will be declared closed and be available for statistical analysis. All data management activities will be done according to the current Standard Operating Procedures (SOPs) of IZKS Mainz.

### 9.4 Storage and archiving of data at the study sites

All study data (patient identification list, source data) and relevant correspondence will be stored in the Investigator Site File (ISF). The ISF, all source data and all relevant documents itemized in section 8 of the ICH Consolidated Guideline on GCP will be archived after end of the study for at least 15 years. Storage and archiving of the electronic data during the study will be assured by the IZKS Mainz. After completion of the study, all electronic data will be handed over to the sponsor.

## 10 ETHICAL AND LEGAL ASPECTS

The procedures set out in this study protocol, pertaining to the conduct, evaluation, and documentation of this study, are designed to ensure that all persons involved in the study abide by good clinical practice (GCP) as applicable and the ethical principles described in the Declaration of Helsinki. The study will be carried out in accordance with local legal and regulatory requirements, particularly regarding the Federal Data Protection Act (BDSG).

### 10.1 Patient information and informed consent

Before being admitted to the clinical study, the patient must consent to participate after being fully informed about the nature, scope, and possible consequences of the study. The documents must be in a language understandable to the patient and must specify who informed the patient. A copy of the signed informed consent document must be given to the patient. The original signed consent document will be retained by the investigator. The investigator will not undertake any study-specific procedures until valid consent has been obtained. If the patient has a primary physician, the investigator should inform the patient’s primary physician about the patient’s participation in the study if the patient agrees to the primary physician being informed.

If the patient is unable to personally date and sign the consent form due to disability, consent must be confirmed orally in the presence of an impartial witness. The witness and the person conducting the informed consent discussions must sign and personally date the consent document.

If the patient is unable to read, oral presentation and explanation of the written informed consent form and information must take place in the presence of an impartial witness. Consent must be confirmed orally and by the personally dated signature of the patient, or by a local legally recognized alternative (e.g., the patient’s thumbprint or mark). The witness and the person conducting the informed consent discussions must also sign and personally date the consent document.

### 10.2 Confidentiality

The name of the patients and other confidential information are subject to medical professional secrecy and the regulations of the Federal Data Protection Act (BDSG). During the clinical study, patients will be identified solely by means of an individual identification code (patient number). Study findings stored on a computer will be stored in accordance with local data protection law and will be handled with strictest confidence. For protection of these data, organizational procedures are implemented to prevent distribution of data to unauthorized persons. The appropriate regulations of data legislation will be fulfilled in its entirety.

The patient will declare in the written consent to release the investigator from the medical professional secrecy to allow identification of patient’s name and/or inspection of original data for monitoring purposes by health authorities and authorized persons (monitors). The investigator will maintain a personal patient identification list (patient numbers with the corresponding patient names) to enable records to be identified.

### 10.3 Responsibilities of principal investigators

The principal investigators of each study site ensure that all persons assisting with the study are adequately informed about the protocol, any amendments to the protocol, the study procedures, and their study-related duties and functions. The principal investigators will maintain a list of appropriately qualified persons to whom they have delegated significant study-related duties (delegation log). Any changes of the study personnel must be communicated without delay to the IZKS Mainz.

### 10.4 Approval of study protocol and substantial amendments

Before the start of the study, the study protocol, informed consent document, and any other appropriate documents are submitted to the independent Ethics Committees (EC). Before the first patient is enrolled in the study, all ethical and legal requirements must be met. The EC will be informed of all subsequent protocol amendments which might require formal approval.

### 10.5 Data Safety and Monitoring Board

An independent Data Safety and Monitoring Board (DSMB) consisting of clinical and statistical experts will be established by the sponsor to review the safety data. The DSMB supervises the progress of the study, monitors the safety data, and gives advice to the sponsor whether to continue, modify, or stop the study. DSMB meetings will be held four times a year.

### 10.6 Insurance

The sponsor takes out a voluntary patient insurance which covers, in its terms and provisions, the legal liability for injuries caused to participating persons and arising out of this research, which is performed strictly in accordance with the scientific protocol as well as with applicable law and professional standards.

The insurance was taken out at HDI-Gerling Industrie Versicherung AG, Niederlassung Düsseldorf, Am Schönenkamp 45, 40599 Düsseldorf. The insurance number is Nr. 57 010315 03015, the maximum limit is € 500.000 per participating person.

Any impairment of health, which might occur in consequence of study participation, must be notified to the insurance company. The patient is responsible for notification. The insured person will agree to all appropriate measures serving for clarification of the cause and the extent of damage as well as the reduction of damage.

The patient is bound to inform the investigator immediately about any treatment by another physician, any adverse events, and any additional drugs taken. The terms and conditions of the insurance must be delivered to the patient, indicating that policy conditions must be observed.

Insurance coverage exists for each patient during the whole participation in the clinical study. Insurance provisions for this clinical study are given in separate agreements.

### 10.7 Agreements

#### 10.7.1 Financing of the study

The study is funded by the sponsor University Hospital Ulm and supported by German Research Foundation (DGF, grant reference number DO 2484_3-1). The general conditions of financing for this study are given in separate agreements.

#### 10.7.2 Report

After conclusion of the study, a report shall be written by the sponsor with the coordinating investigator. The report will include a statistical analysis and an appraisal of the results from a medical viewpoint. It will be based on the items listed in this study protocol.

#### 10.7.3 Publication policy

Any publication of the results, either in part or in total (articles in journals or newspapers, oral presentation, etc.) by the investigators or their representatives require the approval of the sponsor. It is planned to publish the results of the study as an original article in an appropriate medical journal and as presentations at congresses. The publication policy for this study is described in detail in separate agreements.

## 11 SIGNATURES

The present study protocol was subject to critical review and has been approved in the present version by the persons undersigned. The information contained is consistent with

- The current risk-benefit assessment of the study intervention.
- The moral, ethical, and scientific principles governing clinical research as set out in the Declaration of Helsinki and the principles of GCP as applicable.

**Coordinating Investigator/Project Management**

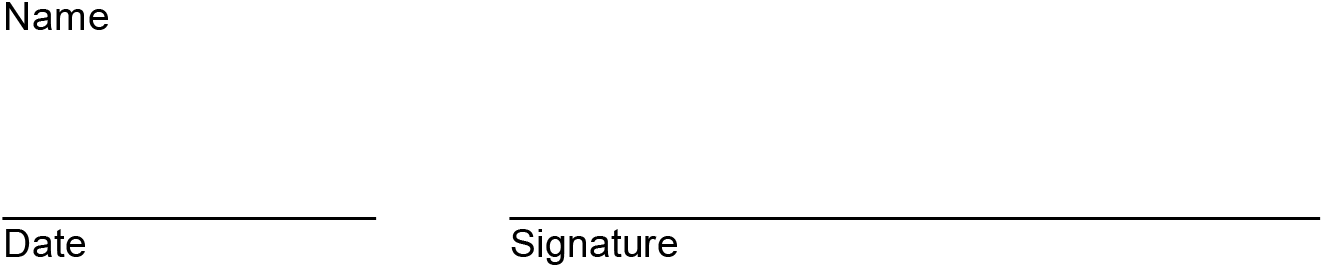

**Biometrician**

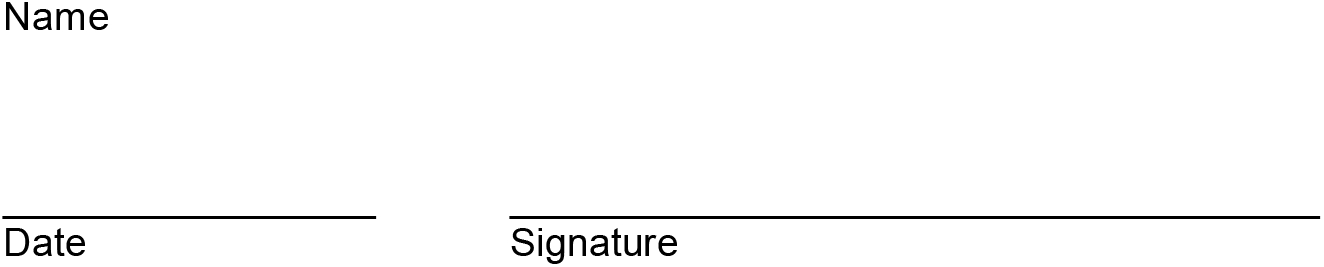

## Data Availability

All data produced in the present study are available upon reasonable request to the authors.

## List of abbreviations

AE: Adverse event
ALS: Amyotrophic lateral sclerosis
ALT: Alanine-Aminotransferase
ALSFRS-R ALS: Functional Rating Scale – Revised
AST: Aspartate-Aminotransferase
BMI: Body Mass Index
CK: Creatinkinase
CNAQ: Council of Nutrition appetite questionnaire
CRF: Case report form
CRP: C-reactive protein
CSF: Cerebospinal fluid
DSMB: Data Safety and Monitoring Board
eCRF: electronic case report form
EQ-5D-5L: Euro Quality of Life-5D-5L
FAS: Full Analysis Set
GCP: Good clinical practice
Hb: Hemoglobin
HCFD: High-caloric fatty diet
Hct: Hematocrit
HDL: High density lipoprotein
ICH: International conference on harmonization of technical requirements for registration of pharmaceuticals for human use
ISF: Investigator site file
ITT: Intention-To-Treat
IZKS: Interdisciplinary Center for Clinical Trials
LDL: Low density lipoprotein
MedDRA: Medical dictionary for regulatory activities
PPS: Per-protocol analysis set
RBC: Red blood cells
ROADS: Rasch-Built Overall ALS Disability Scale
SAE: Serious adverse event
SAP: Statistical analysis plan
SOP: Standard operating procedure
SVC: Slow vital capacity
UFD: Ultra-high-caloric fatty diet
UNQ: Ulm Nutrition Questionnaire
WBC: White Blood Cells

## REFERENCES

1 Brooks BR, Miller RG, Swash M, Munsat TL. El Escorial revisited: revised criteria for the diagnosis of amyotrophic lateral sclerosis. Amyotrophic lateral sclerosis and other motor neuron disorders : official publication of the World Federation of Neurology, Research Group on Motor Neuron Diseases 2000; 1(5): 293–9.

2 Cedarbaum JM, Stambler N, Malta E, et al. The ALSFRS-R: a revised ALS functional rating scale that incorporates assessments of respiratory function. BDNF ALS Study Group (Phase III). Journal of the neurological sciences 1999; 169(1-2): 13–21.

3 Fournier CN, Bedlack R, Quinn C, et al. Development and Validation of the Rasch-Built Overall Amyotrophic Lateral Sclerosis Disability Scale (ROADS). JAMA neurology 2019; 30(2758019).

4 Longinetti E, Regodón Wallin A, Samuelsson K, et al. The Swedish motor neuron disease quality registry. Amyotrophic lateral sclerosis & frontotemporal degeneration 2018; 19(7-8): 528–37.

5 Alonso A, Hernán MA. Temporal trends in the incidence of multiple sclerosis: a systematic review. Neurology 2008; 71(2): 129–35.

6 Larson TC, Kaye W, Mehta P, Horton DK. Amyotrophic Lateral Sclerosis Mortality in the United States, 2011-2014. Neuroepidemiology 2018; 51(1-2): 96–103.

7 Global, regional, and national burden of motor neuron diseases 1990-2016: a systematic analysis for the Global Burden of Disease Study 2016. Lancet neurology 2018; 17(12): 1083–97.

8 Brettschneider J, Del Tredici K, Toledo JB, et al. Stages of pTDP-43 pathology in amyotrophic lateral sclerosis. Annals of neurology 2013.

9 Braak H, Brettschneider J, Ludolph AC, Lee VM, Trojanowski JQ, Del Tredici K. Amyotrophic lateral sclerosis--a model of corticofugal axonal spread. Nature reviews Neurology 2013; 9(12): 708–14.

10 Bensimon G, Doble A. The tolerability of riluzole in the treatment of patients with amyotrophic lateral sclerosis. Expert opinion on drug safety 2004; 3(6): 525–34.

11 EdavaroneStudyGroup. Safety and efficacy of edaravone in well defined patients with amyotrophic lateral sclerosis: a randomised, double-blind, placebo-controlled trial. Lancet neurology 2017; 15(17): 30115–1.

12 Miller TM, Pestronk A, David W, et al. An antisense oligonucleotide against SOD1 delivered intrathecally for patients with SOD1 familial amyotrophic lateral sclerosis: a phase 1, randomised, first-in-man study. Lancet neurology 2013; 12(5): 435–42.

13 Miller T, Cudkowicz M. Results from the Phase 3 VALOR study and its open-label extension: evaluating the clinical efficacy and safety of tofersen in adults with ALS and confirmed SOD1 mutation. American Neurological Association Annual Meeting 2021.

14 Desport JC, Preux PM, Magy L, et al. Factors correlated with hypermetabolism in patients with amyotrophic lateral sclerosis. The American journal of clinical nutrition 2001; 74(3): 328–34.

15 Gorges M, Vercruysse P, Muller HP, et al. Hypothalamic atrophy is related to body mass index and age at onset in amyotrophic lateral sclerosis. Journal of neurology, neurosurgery, and psychiatry 2017; 8(315795): 2017–315795.

16 Vercruysse P, Sinniger J, El Oussini H, et al. Alterations in the hypothalamic melanocortin pathway in amyotrophic lateral sclerosis. Brain : a journal of neurology 2016; 139(Pt 4): 1106–22.

17 Desport JC, Preux PM, Truong TC, Vallat JM, Sautereau D, Couratier P. Nutritional status is a prognostic factor for survival in ALS patients. Neurology 1999; 53(5): 1059–63.

18 Marin B, Desport JC, Kajeu P, et al. Alteration of nutritional status at diagnosis is a prognostic factor for survival of amyotrophic lateral sclerosis patients. Journal of neurology, neurosurgery, and psychiatry 2011; 82(6): 628–34.

19 Dupuis L, Oudart H, Rene F, Gonzalez de Aguilar JL, Loeffler JP. Evidence for defective energy homeostasis in amyotrophic lateral sclerosis: benefit of a high-energy diet in a transgenic mouse model. Proceedings of the National Academy of Sciences of the United States of America 2004; 101(30): 11159–64.

20 Dupuis L, Corcia P, Fergani A, et al. Dyslipidemia is a protective factor in amyotrophic lateral sclerosis. Neurology 2008; 70(13): 1004–9.

21 Dorst J, Kuhnlein P, Hendrich C, Kassubek J, Sperfeld AD, Ludolph AC. Patients with elevated triglyceride and cholesterol serum levels have a prolonged survival in amyotrophic lateral sclerosis. Journal of neurology 2011; 258(4): 613–7.

22 Michels S, Kurz D, Rosenbohm A, et al. Association of blood lipids with onset and prognosis of amyotrophic lateral sclerosis: results from the ALS Swabia registry. Journal of neurology 2023; 28(10): 023–11630.

23 Wills AM, Hubbard J, Macklin EA, et al. Hypercaloric enteral nutrition in patients with amyotrophic lateral sclerosis: a randomised, double-blind, placebo-controlled phase 2 trial. Lancet 2014; 383(9934): 2065–72.

24 Ludolph AC, Dorst J, Dreyhaupt J, et al. Effect of high-caloric nutrition on survival in amyotrophic lateral sclerosis. Annals of neurology 2019; 17(10): 25661.

25 Dorst J, Schuster J, Dreyhaupt J, et al. Effect of high-caloric nutrition on serum neurofilament light chain levels in amyotrophic lateral sclerosis. Journal of neurology, neurosurgery, and psychiatry 2020; 91(9): 1007–9.

26 Steinacker P, Feneberg E, Weishaupt J, et al. Neurofilaments in the diagnosis of motoneuron diseases: a prospective study on 455 patients. Journal of neurology, neurosurgery, and psychiatry 2016; 87(1): 12–20.

27 Verde F, Steinacker P, Weishaupt JH, et al. Neurofilament light chain in serum for the diagnosis of amyotrophic lateral sclerosis. Journal of neurology, neurosurgery, and psychiatry 2019; 90(2): 157–64.

28 Dorst J, Doenz J, Kandler K, et al. Fat-rich versus carbohydrate-rich nutrition in ALS: a randomised controlled study. Journal of neurology, neurosurgery, and psychiatry 2022; 93(3): 298–302.

29 Dorst J, Dupuis L, Petri S, et al. Percutaneous endoscopic gastrostomy in amyotrophic lateral sclerosis: a prospective observational study. Journal of neurology 2015.

30 Witzel S, Frauhammer F, Steinacker P, et al. Neurofilament light and heterogeneity of disease progression in amyotrophic lateral sclerosis: development and validation of a prediction model to improve interventional trials. Transl Neurodegener 2021; 10(1): 021–00257.

31 Ludolph AC, Schuster J, Dorst J, et al. Safety and efficacy of rasagiline as an add-on therapy to riluzole in patients with amyotrophic lateral sclerosis: a randomised, double-blind, parallel-group, placebo-controlled, phase 2 trial. Lancet neurology 2018; 18(18): 30176–5.

32 Dupuis L, Dengler R, Heneka MT, et al. A randomized, double blind, placebo-controlled trial of pioglitazone in combination with riluzole in amyotrophic lateral sclerosis. PloS one 2012; 7(6): e37885.

33 Rohlmann F MR, Goldschmidt L. Randomisation program ROM. DVMD Conference 2004M; see Schweizer, B et al (ed): Dokumentation - der Schritt ins 3 Jahrtausend 8 DVMD-Tagung in Ulm, 1 - 2 April 2004 Universitätsverlag Ulm 2004 2004.

